# Evolution of Service Metrics and Utilisation of Evidence-Based Outcome Measures in Anterior Cruciate Ligament Reconstruction Rehabilitation: An Observational Review of Two Cohorts in a Public Hospital Physiotherapy Department

**DOI:** 10.1101/2020.03.26.20044032

**Authors:** Kirby Tuckerman, Wendy Potts, Milad Ebrahimi, Corey Scholes, Mark Nelson

## Abstract

**Objective:** Determine in patients undergoing supervised rehabilitation post ACL reconstruction in a public hospital, whether a new model of care incorporating a phase-based program, compared to standard care, increased physiotherapist utilisation of outcome measures, improved service metrics such as attendance and rehabilitation completion rates, as well as increased self-reported knee function and activity levels.

**Methods:** Patients attending outpatient physiotherapy after ACL reconstruction at a metropolitan public hospital (N = 132) were included in retrospective chart review to assess utilisation of outcomes such as quadriceps and hamstrings strength assessment, patient attendance and rehabilitation completion. Phone followup (minimum one year) was conducted to retrieve patient-reported measures of knee function (IKDC) and activity (Tegner Activity Scale). Patients were categorised by rehabilitation model of care (contemporary - time based [N = 93] vs new - phase based [N = 39]) and logistic regression used to assess the influence of patient factors and model of care on outcomes.

**Results:** Patients included for analysis were aged 25 years at surgery (IQR 20.3 - 30.8), with 42.4% of non-Australia country of origin. Compliance was equivalent between models of care and completion rates (formally discharged by therapist) were low (30-38%). The probability of a patient receiving strength assessment was significantly associated with model of care, sex, BMI and the number of sessions attended. The probability of a patient being recorded as discharged from the program was significantly associated with the model of care, as well as the duration and number of sessions.

**Conclusion:** The transition to a new model of care incorporating a phase-based rehabilitation program increased physiotherapist utilisation of certain evidence-based outcome measures, increased total duration of rehabilitation and increased the total number of sessions attended. Despite this, rehabilitation completion rates remained low, and no change was demonstrated with respect to self-reported knee function and activity levels.

**Level of evidence:** III, retrospective case-control study

## Introduction

Anterior cruciate ligament (ACL) rupture is a common injury that usually occurs during non-contact pivoting or twisting movements of the knee ^19,21^. A ruptured ACL can result in instability and reduced functional abilities including high level sports ^17,28^. Additionally, it leads to an increased risk of developing knee osteoarthritis ^28^. Management of ACL rupture aims to restore stability and optimise patient function. Management approaches can be either conservative (exercise-based rehabilitation programs) or surgical (ACL reconstruction) (ACLR)^20^. Regardless of management approach, ACL ruptures require prolonged rehabilitation to regain functional abilities. While the importance of ACL rehabilitation is widely accepted in the literature ^5,16,19,26^, specific rehabilitation practices differ.

Variation in rehabilitation exists with respect to setting, progression, duration and return to functional activities and sport ^8,15,16^ The literature suggests ACLR rehabilitation should address several factors including range of motion (ROM), muscle function (neuromuscular control, strength, endurance, power), performance and psychological factors ^8,19,23,25^. Additionally, there is good evidence that rehabilitation should be progressive in nature and based on the achievement of objective outcomes and a battery of tests used to determine readiness for discharge and return to sport ^5,8,16,19,23^. Suggested tests include strength measurement of several muscle groups, hop tests and movement quality assessment. Despite these recommendations, existing literature demonstrates a disparity in discharge criteria used ^2,4,13^. It must also be recognised that there is a significant risk of re-injury post ACLR, with reported re-rupture rates from 5%-23% ^8^, and risk increasing in those returning to sport too early ^19,28^.

The optimal length of supervised rehabilitation post ACLR is currently unclear due to a lack of high-quality studies ^9,19^. Literature suggests many ACLR patients receive insufficient rehabilitation due to early discharge from services or inadequate content of rehabilitation ^16^. The level of individual physiotherapists’ experience and knowledge regarding evidence-based recommendations varies, and when considering the public healthcare setting, rehabilitation services are provided with finite resources. The aim of this study was to determine in patients electing to undergo supervised rehabilitation in a physiotherapy department in a public hospital, whether a new model of care incorporating a phase-based program, compared to standard care provided the following benefits:

- Increased physiotherapist utilisation of evidence-based outcome measures
- Improved service metrics such as attendance and rehabilitation completion rates
- Increased self-reported knee function and activity levels

## Methods

An observational cohort study with historical control was used to review service metrics and utilisation of evidence-based outcome measures before and after a change in model of care for ACLR rehabilitation. Ethical approval was granted by the Metro South human research ethics committee (HREC/16/QPAH/732) prior to patient screening and data retrieval.

A list of patients who underwent primary ACLR at a metropolitan public hospital, between November 2014 and December 2017 was exported from ORMIS/CERNER by hospital administration (N=262). Procedures coded as revision surgery (N=3) and surgeries outside the specified date range (N=35) were removed.

During the chart review process, revision ACLR procedures, a primary ACLR with concurrent surgical treatment of other ligaments (PCL, LCL, MCL, PLC) and those referred to an external physiotherapy service provider on discharge from the ward were excluded (N=92). The final list included 132 patients who had undergone ACLR and rehabilitation at the hospital outpatient physiotherapy department. The final 132 patients were split into two groups using a surgery date of 1st of November 2016 as the group cut off, which aimed to ensure the majority of the patients who had surgery after this date underwent rehabilitation using the new model of care (CONTROL N=93, NEW N=39).

### Clinical practice prior to model of care change (CONTROL)

Following ACLR, patients were referred to the hospital physiotherapy outpatient department for an initial appointment within one to two weeks of their surgery. Appointments were thirty minutes in duration. Follow-up appointments were completed weekly for the initial postoperative period and then at less frequent intervals depending on the treating clinicians clinical judgement. Rehabilitation was a 1:1 format and guided by the individual clinician’s knowledge and clinical preferences regarding ACLR rehabilitation. During rehabilitation, patients were often assessed and treated by a number of physiotherapists with a range of experience levels.

### Model of care change: Phase-based rehabilitation program (NEW)

A review of ACLR rehabilitation practice in the physiotherapy department occurred in September 2016. A new model of care was developed aiming to provide a pathway to consistently deliver evidence-based care and enhance patient outcomes. A four-phase outcome-based program was developed based on current evidence, particularly the Randall Cooper ACL Rehabilitation Guide ^7^. Phase progression and discharge (return to function and sport) was based on outcome measure performance (Appendix 1). A progressive exercise program corresponding to each phase was developed. Patients were invited to participate in a combination of 1:1 and group sessions held at similar intervals to the CONTROL group. Both individual and group classes followed the same packaged model of care (Appendix 2), with the aim of improving patients’ understanding of the rehabilitation process and increasing motivation and adherence.

### Data collection

#### Chart Audits

Pilot chart audits were completed on 50 randomly selected records to investigate availability and quality of data. Following screening for eligibility, 23 CONTROL and 2 NEW patient records were collected using a custom data collection tool (Excel, Microsoft, USA), which was adjusted following feedback from the pilot review. All time-related data was referenced from the date of surgery. After the data collection tool was finalised, the pilot charts were reviewed again using the finalised tool. Data collection for the remaining 82 patient records was then undertaken by a single physiotherapist to negate the potential for limited inter-observer reliability.

#### Phone calls

Follow-up phone calls were made between one- and three-years post-surgery to all patients who had undergone an ACLR and had received some form of rehabilitation at the physiotherapy outpatient department. Patients who completed rehabilitation, were removed due to lack of attendance or withdrew during the rehabilitation process were contacted by a single physiotherapist using phone numbers provided by hospital administration. Data was collected regarding patient’s recollection of their rehabilitation and return to activity and sport. Reasons for patients failing to complete rehabilitation was also explored. Patients were sent a text advising them the research team would be attempting to call and included a link to a web-based form (Survey Monkey) asking their preferred time to be contacted. After three unsuccessful attempts to contact a patient, a web-based form (Survey Monkey) was communicated via text message for patients to complete in their own time.

#### Outcomes

‘Physiotherapist adherence to evidence-based outcome measures’ and ‘service metrics’ were collected via the chart audits.

The following outcome measures were recorded as assessed or not assessed (yes or no):

- Quadriceps strength (1 repetition max (1RM) leg press, manual muscle test (MMT) or hand-held dynamometry (HHD))
- Hamstring strength (MMT or HHD)
- Kinetic chain strength (Calf, glutes, trunk)
- Hop tests (single leg hop for distance or triple crossover hop)
- Balance (Star excursion balance test (SEBT) or single leg balance)
- Knee range of motion (ROM)
- Time to return to running (weeks)

The following service metrics were also collected:

- Number of individual sessions attended
- Number of group sessions attended
- Number of failed attendances
- Total occasions of service
- Rehabilitation completion status
- Length of physiotherapy input (weeks)
- Number of clinicians

#### Patient reported outcomes

IKDC subjective scores, Tegner activity scores and ACL rerupture rates were collected via follow-up phone calls. The IKDC is an 18 question evaluation that measures symptoms (7 items), activities of daily living (9 items) and sport (1 item), and comparative knee function (1 item - not included in total score).

### Data and Statistical Analysis

The completed chart review spreadsheet (Excel, Microsoft, USA) and patient followup response sheet (Sheets, Google, USA) were transferred into Matlab (Mathworks, USA) and linked by patient unique identifier (person-level linkage) into one combined dataset for analysis in a statistical software package (Minitab, Minitab Inc, USA). Categorical data was recoded to standardize spelling variations or for reasons listed (**Table 1**). Continuous variables were assessed for normality using Anderson-Darling tests. Patient characteristics were summarised using median and interquartile range for continuous variables and proportions for categorical variables. Patient demographics, service utlisation and patient outcomes were compared between groups using unadjusted Mann-whitney U tests for unmatched comparisons of continuous variables and *X*^2^ analysis with likelihood ratio for categorical variables with >2 responses, or Fisher Exact test otherwise. Attendance ratios were calculated between the total number of sessions labelled *failed to attend* relative to the total number of sessions for each group and compared with Fisher’s exact test. Loss of knee extension was defined as 5° or greater fixed flexion angle and the proportion of positive (>5°) measurements were compared between groups with Fisher’s exact test. Backward stepwise binary logistic regression models were constructed to compare service metrics (quadriceps and hamstrings assessment) and rehabilitation status (completed, not completed). A similar model was constructed for ipsilateral rerupture incidence with a weighting vector included to compensate for unbalanced proportions between response categories. Alpha for univariate analyses was set at 0.05, while model alpha for variable inclusion was set at 0.15. Model fit was assessed with adjusted R^2^ (%) and effect sizes expressed with odds ratio with 95% confidence intervals.

**Table 1:** Data recoding

| **Variable** | **Original responses** | **Recoded responses** | **Rationale** |
| --- | --- | --- | --- |
| Country of Origin | Australia<br>New Zealand<br>South Africa<br>England<br>Phillipines<br>Fiji<br>India<br>China | Australia<br>Asia-Pacific<br>Other | Prevent quasi-separation in between-group comparisons |
| Comorbidities | Free - text | None<br>Single<br>Multiple | Include as a model predictor |
| Secondary diagnosis | Free text | Medial meniscus pathology (Yes; No)<br>Lateral meniscus pathology (Yes; No) | Include as a model predictor |
| Quadriceps measurement | 1RM<br>Not recorded<br>HHD<br>MMT | Yes<br>No | Prevent quasi-separation in between-group comparisons; address missing data |
| Hamstrings measurement | 1RM<br>Not recorded<br>HHD<br>MMT | Yes<br>No | Prevent quasi-separation in between-group comparisons; address missing data |
| Balance measurement | SL Balance<br>SEBT<br>Not recorded | Yes<br>No | Prevent quasi-separation in between-group comparisons; address missing data |
| Rehab Status | Completed (Discharged)<br>Failed to attend<br>Withdrew | Completed<br>Did not complete | Prevent quasi-separation in between-group comparisons; |
| Number of staff involved | Integer 1 - 9 | 1-3<br>4-6<br>7+ | Prevent quasi-separation in between-group comparisons |
| Complications | Free text | Yes<br>No | Include as a model predictor |
| Knee extension angle | Continuous (Degrees) | $\geq 5$<br><5 | Relate to a clinically meaningful threshold |
| Reason for failed to attend | Free text | Too hard to attend<br>Discharge belief<br>Happy with knee<br>Changed service | Summarise into key themes |
| IKDC - subjective knee function score | Continuous, 0 - 100 | ≥ PASS<br>< PASS | Relate to a clinically meaningful threshold |

## Results

### Patient characteristics

A sample of 132 patients were identified (combined median age 25 [IQR 20.3 - 30.8yrs]; BMI 26.1 [23.7 -28.5kg/m^2^]; 64.4% male; 42.4% Non-Australian country of origin) meeting inclusion criteria for analysis (**Figure 1**). The sample was split into two groups based on surgery date, including conventional model of care group (CONTROL, N = 93) and the new model of care (NEW, N = 39). The two groups were equivalent for baseline characteristics, except for the proportion of females (48.7% NEW, 30.1% CONTROL (P =0.04)) (**Table 2**).

**Table 2:** Baseline characteristics and initial evaluation of the patient groups separated by the ACL model of care change

|  | NEW (N = 39) | CON (N = 93) | P - value |
| --- | --- | --- | --- |
| Age (years) | 24 (19 - 31) | 25 (21 - 30.5) | 0.51 |
| Female (%) | 48.7 | 30.1 | **0.04** |
| BMI (kg/m<sup>2</sup>) | 26.8 (23.7 - 31.5) | 25.4 (23.6 - 27.8) | 0.16 |
| Injury to Surgery (weeks) | 26.4 (16.1 - 84.9) | 30.4 (18 - 58.7) | 0.98 |
| Surgery - Initial Appt (weeks) | 1.4 (1.1 - 2.6) | 1.6 (1.1 - 2.1) | 0.61 |
| Country of origin (%) |  |  |  |
| Australia | 59 | 57 | 0.95 |
| Asia-Pacific | 23.1 | 22.6 |  |
| *Other* | 18 | 20.4 |  |
| Contralateral Injury (%) | 10.3 | 12.9 | 0.67 |
| Meniscal Injury (%) |  |  |  |
| *Medial* | 30.8 | 40.9 | 0.27 |
| *Lateral* | 46.2 | 32.3 | 0.13 |
| Comorbidities (%) |  |  |  |
| *Single* | 41 | 36.7 | 0.89 |
| *Multiple* | 15.4 | 16.1 |  |
| Prehabilitation |  |  |  |
| *Yes* | 61.5 | 55.9 | 0.79 |
| *No* | 5.1 | 7.5 |  |
| *Not recorded* | 33.3 | 36.6 |  |
| Prescribed Weightbearing |  |  |  |
| *Full* | 69.2 | 58.1 | 0.10 |
| *Partial* | 20.5 | 37.6 |  |
| *Non* | 10.3 | 4.3 |  |
| Prescribed Brace | 46.2 | 57 | 0.26 |
| Range of motion restriction | 38.5 | 41.9 | 0.71 |
| Pain Level (NRS) | 3 (2 - 5.8) | 4 (3 - 6) | 0.40 |
| Loss of extension (%) | 13.5 | 22 | 0.27 |

**Figure 1:**
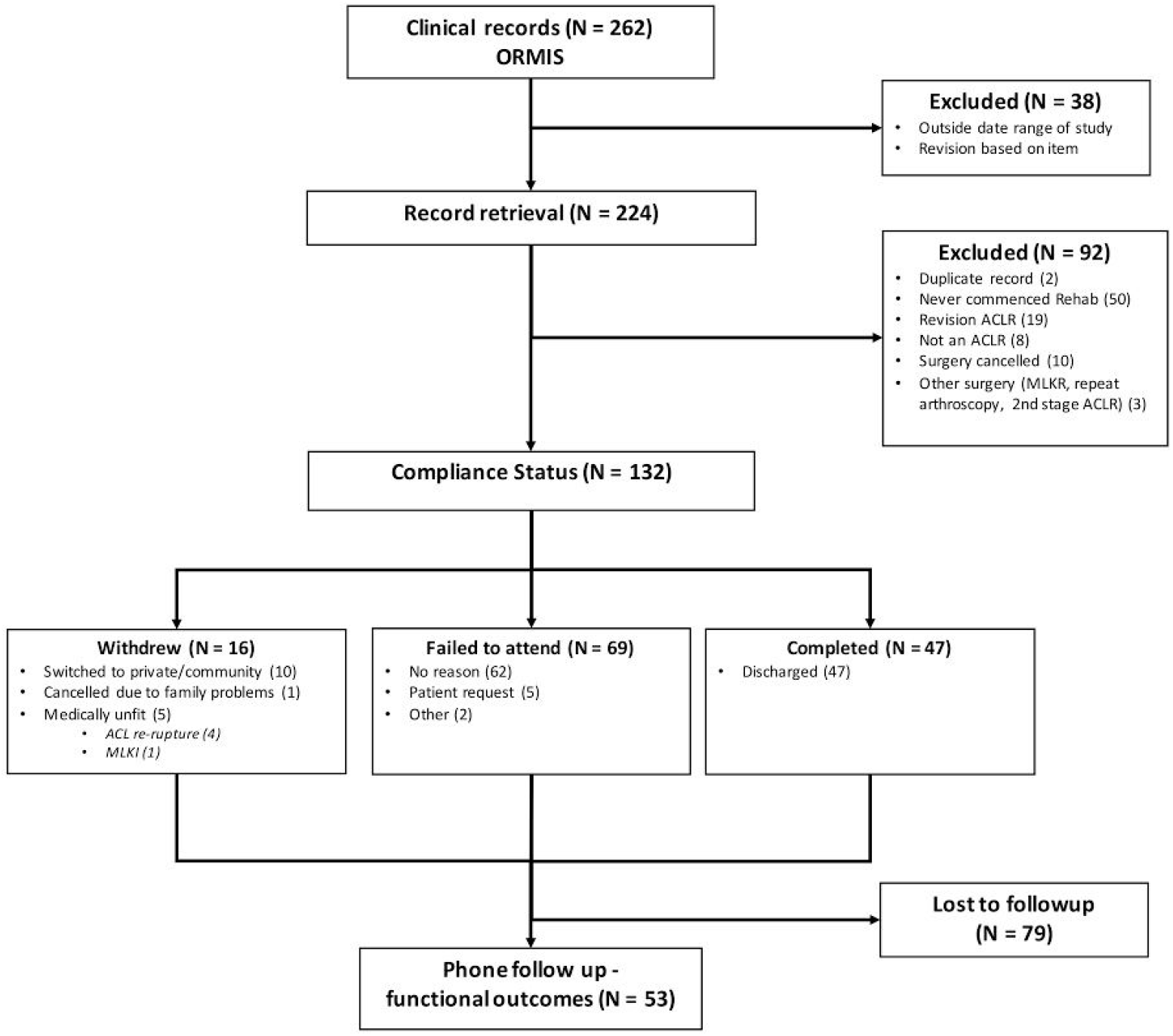
STROBE ^24^ flow diagram of screening and analysis of patients in the study. MKLI, multiligament knee injury; MLKR, multiligament knee reconstruction; ORMIS, operating room management information system.

### Service Utilisation

The total number of physiotherapy sessions attended (11 vs. 8, p=0.02) and the duration (weeks) of physiotherapy input (36.8 vs. 23.6, p=0.01; **Table 3**) were the only service metrics with a significant difference between groups. The percentage of patients who completed rehabilitation and were discharged from the physiotherapy service was 30.8% NEW compared to 37.6% CONTROL (p=0.61). Patients discharged due to a failure to attend appointments was 59% NEW compared to 49.5% CONTROL, and 10.3% NEW compared to 12.9% CONTROL (p=0.61) withdrew from physiotherapy (**Table 3**). Voluntary withdrawal from rehabilitation was dominated by patients changing rehabilitation facilities (**Figure 2**). Patient reasons for reduced attendance was investigated in follow-up phone calls (N=53), with 39% stating they believed they had been discharged, while 32% found it too hard to attend (**Figure 3)**. The number of physiotherapists involved in patient care was not significantly different between groups, with greater than four physiotherapists involved in 64.1% of cases in NEW group compared to 46.2% in CONTROL (**Table 3**). Physiotherapy was supplemented by access to gym or exercise equipment in more than 50% of cases in both groups (**Table 3**). There was no significant difference in physiotherapy prior to surgery or ‘prehabilitation’ with 61.5% NEW compared to 55.9% in CONTROL and participation unknown in up to 36% of participants across the groups.

**Table 3:** Comparison of Service Utilisation between groups

|  | NEW (N = 39) | CON (N = 93) | P value |
| --- | --- | --- | --- |
| Completion status (%) |  |  |  |
| *Completed (discharged)* | 30.8 | 37.6 | 0.61 |
| *Failed to attend* | 59 | 49.5 |  |
| *Withdrew* | 10.3 | 12.9 |  |
| Compliance (%) | 23 | 20.4 | 0.24 |
| Sessions attended (N) | 11 (8 - 15) | 8 (4 - 12) | **0.02** |
| Duration (weeks) | 36.8 (20.5 - 44.5) | 23.6 (9.5 - 36.9) | **0.01** |
| Staff heterogeneity |  |  |  |
| *1-3* | 35.9 | 53.8 | 0.15 |
| *4-6* | 48.7 | 37.6 |  |
| *7+* | 15.4 | 8.6 |  |
| Rehabilitation supplementation |  |  |  |
| *Yes* | 59 | 54.8 | 0.56 |
| *No* | 5.1 | 10.8 |  |
| *Not recorded* | 35.9 | 34.4 |  |

**Figure 2:**
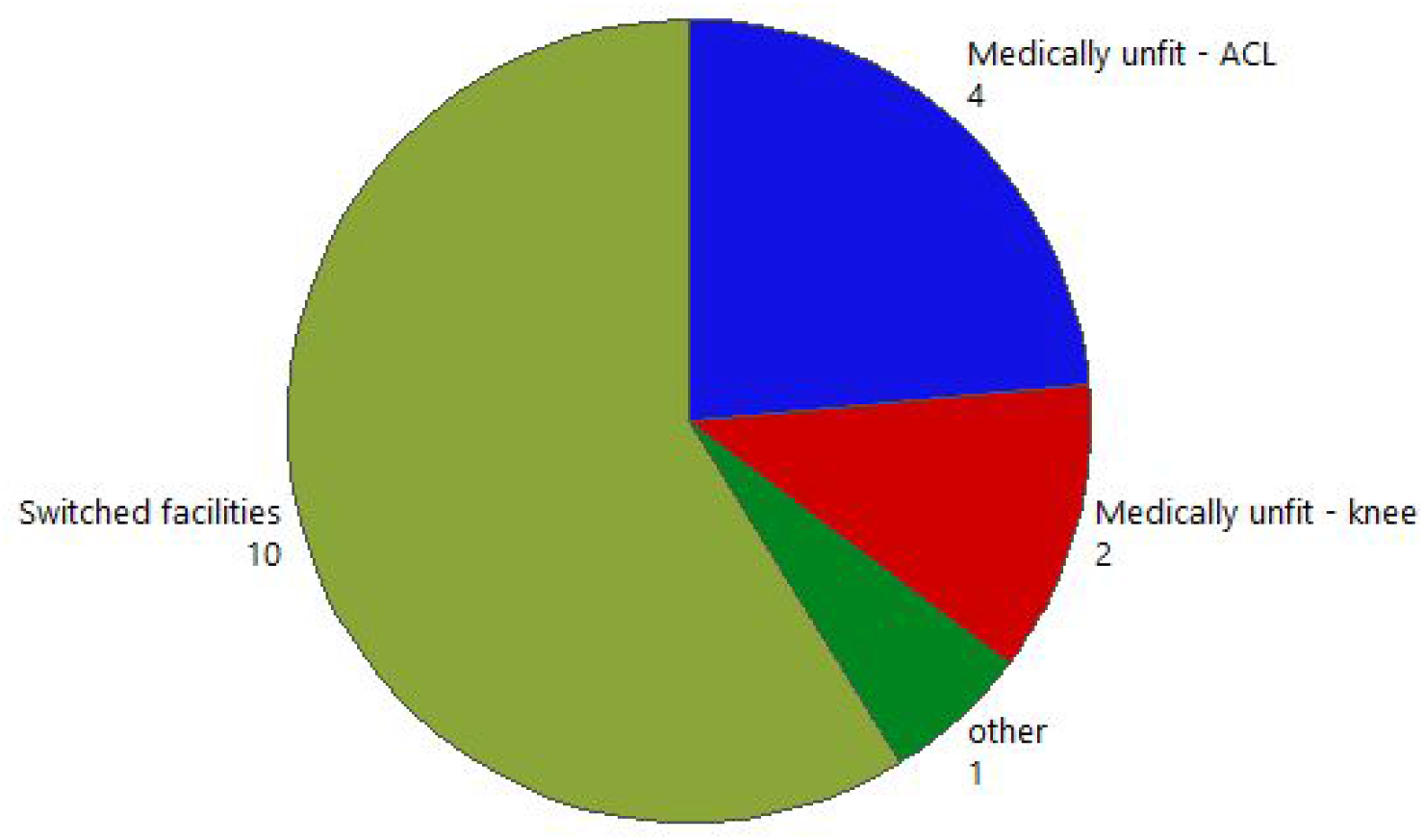
Reason for withdrawal from postoperative rehabilitation (labels are counts)

**Figure 3:**
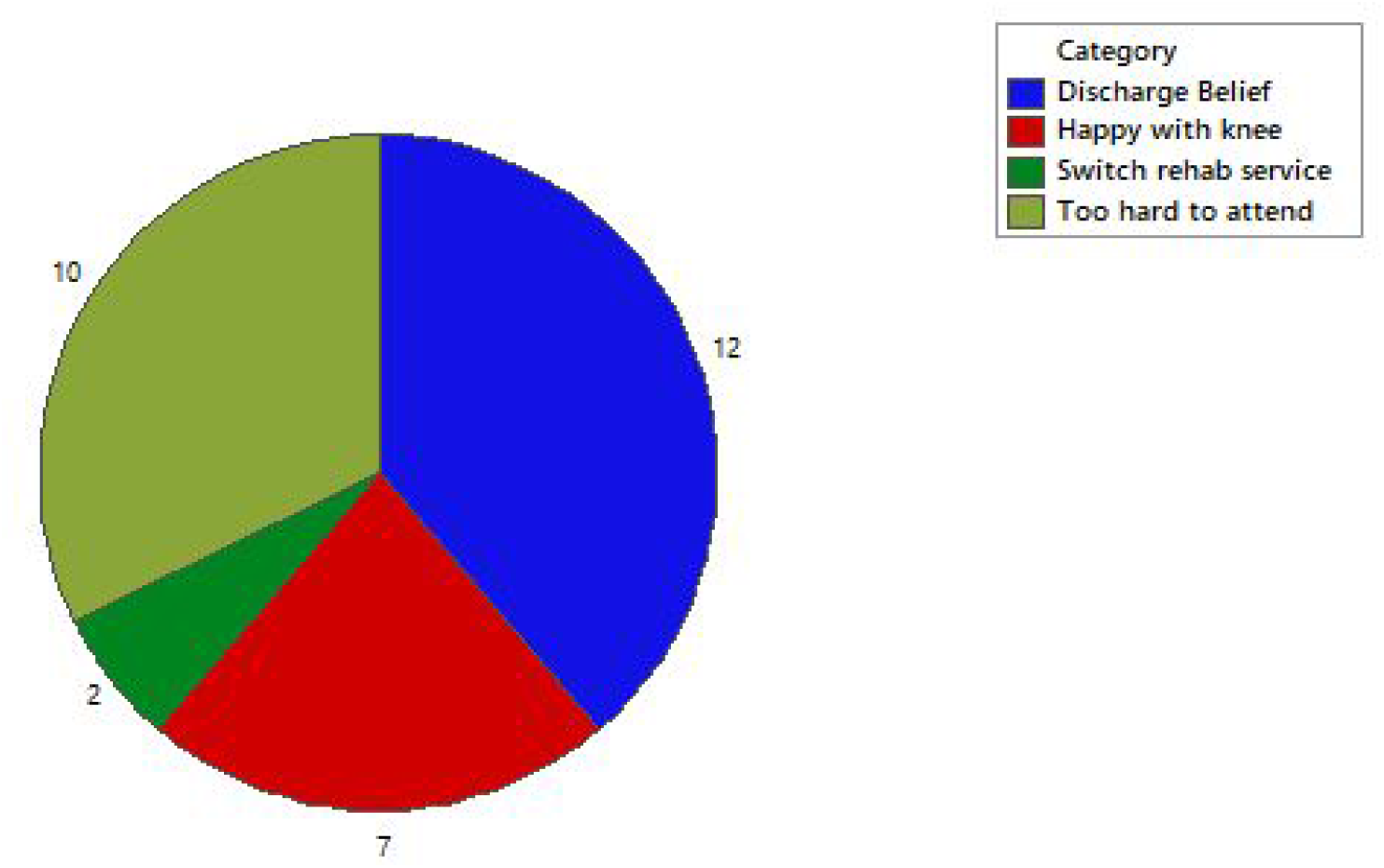
Reason for failure to attend. Labels are counts

### Physiotherapist use of evidence-based outcome measures

The NEW model of care was associated with significantly higher rates of assessment of strength of the quadriceps, the muscles of the kinetic chain and the quality of single leg squat performance (**Table 4**). The assessment of hamstring strength, balance and hop testing all increased following the model of care change but this was not statistically significant between groups. The NEW group took an average of six weeks longer to commence running following initial physiotherapy appointment, starting after 21.1 weeks (17.4-24.9) compared to 15.9 (12-19.3) weeks for CONTROL (**Table 5**).

**Table 4:** Comparison between groups of outcome measure assessment incidence (%) during rehabilitation

|  | NEW (N = 39) | CON (N = 93) | P value |
| --- | --- | --- | --- |
| **Quadriceps strength** | 84.6 | 63.4 | **0.01** |
| **Hamstrings strength** | 56.4 | 49.5 | 0.47 |
| **Kinetic chain assessment** | 84.6 | 41.8 | **<0.001** |
| **Single leg squat assessment** | 87.2 | 61.5 | **0.002** |
| **Hop test** | 59 | 42.9 | 0.09 |
| **Balance** | 71.8 | 70.3 | 0.87 |

**Table 5:**
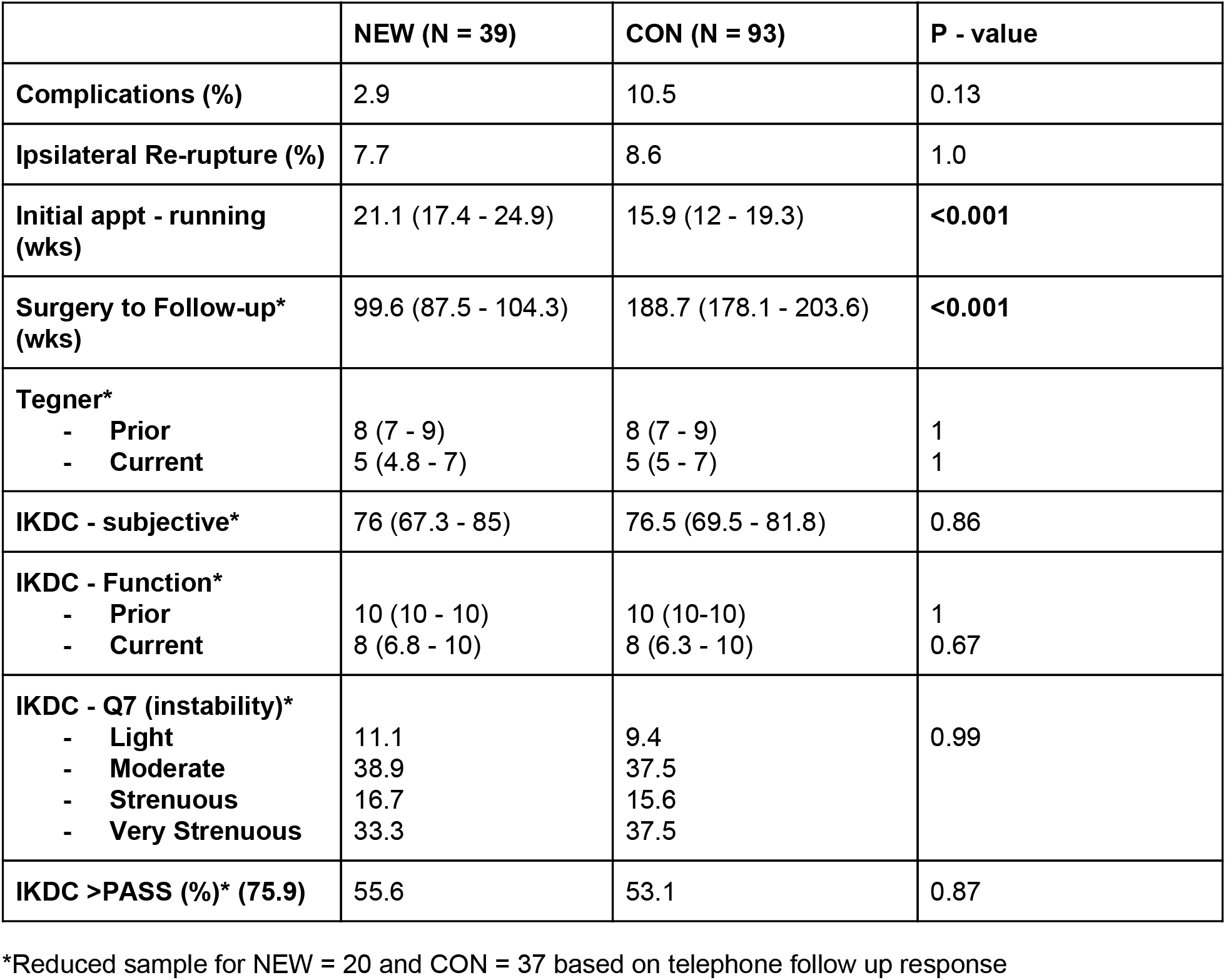
Comparison between groups for patient outcomes during and following rehabilitation.

|  | **NEW (N = 39)** | **CON (N = 93)** | **P - value** |
| --- | --- | --- | --- |
| **Complications (%)** | 2.9 | 10.5 | 0.13 |
| **Ipsilateral Re-rupture (%)** | 7.7 | 8.6 | 1.0 |
| **Initial appt - running (wks)** | 21.1 (17.4 - 24.9) | 15.9 (12 - 19.3) | **<0.001** |
| **Surgery to Follow-up\* (wks)** | 99.6 (87.5 - 104.3) | 188.7 (178.1 - 203.6) | **<0.001** |
| **Tegner\*** |  |  |  |
| - **Prior** | 8 (7 - 9) | 8 (7 - 9) | 1 |
| - **Current** | 5 (4.8 - 7) | 5 (5 - 7) | 1 |
| **IKDC - subjective\*** | 76 (67.3 - 85) | 76.5 (69.5 - 81.8) | 0.86 |
| **IKDC - Function\*** |  |  |  |
| - **Prior** | 10 (10 - 10) | 10 (10-10) | 1 |
| - **Current** | 8 (6.8 - 10) | 8 (6.3 - 10) | 0.67 |
| **IKDC - Q7 (instability)\*** |  |  |  |
| - **Light** | 11.1 | 9.4 | 0.99 |
| - **Moderate** | 38.9 | 37.5 |  |
| - **Strenuous** | 16.7 | 15.6 |  |
| - **Very Strenuous** | 33.3 | 37.5 |  |
| **IKDC >PASS (%)\* (75.9)** | 55.6 | 53.1 | 0.87 |

\*Reduced sample for NEW = 20 and CON = 37 based on telephone follow up response

### Long term outcomes

There was no difference in Tegner Activity Scales between groups, with an average score of 8 prior to ACL rupture and 5 at follow-up. The average IKDC-Function score was consistent across both groups with 10 prior to ACL rupture and 8 at follow-up. IKDC-SKF Patient Acceptable Symptom State (PASS) was above the recommended threshold score of 75.9 ^22^ in 55.6% of participants in NEW group and 53.1% in CONTROL group.

### Logistic Regressions

The probability of a quadriceps strength assessment during rehabilitation was associated with longer physiotherapy duration and the NEW model of care (**Table 6**). Those with higher BMI were less likely to undergo hamstrings assessment, as were females. Attending more sessions was positively associated with the probability of hamstring assessment (**Table 6**). The probability of a patient completing rehabilitation to discharge was significantly associated with a longer duration of physiotherapy, an increased number of attended sessions, and the CONTROL model of care (**Table 6**).

**Table 6:**
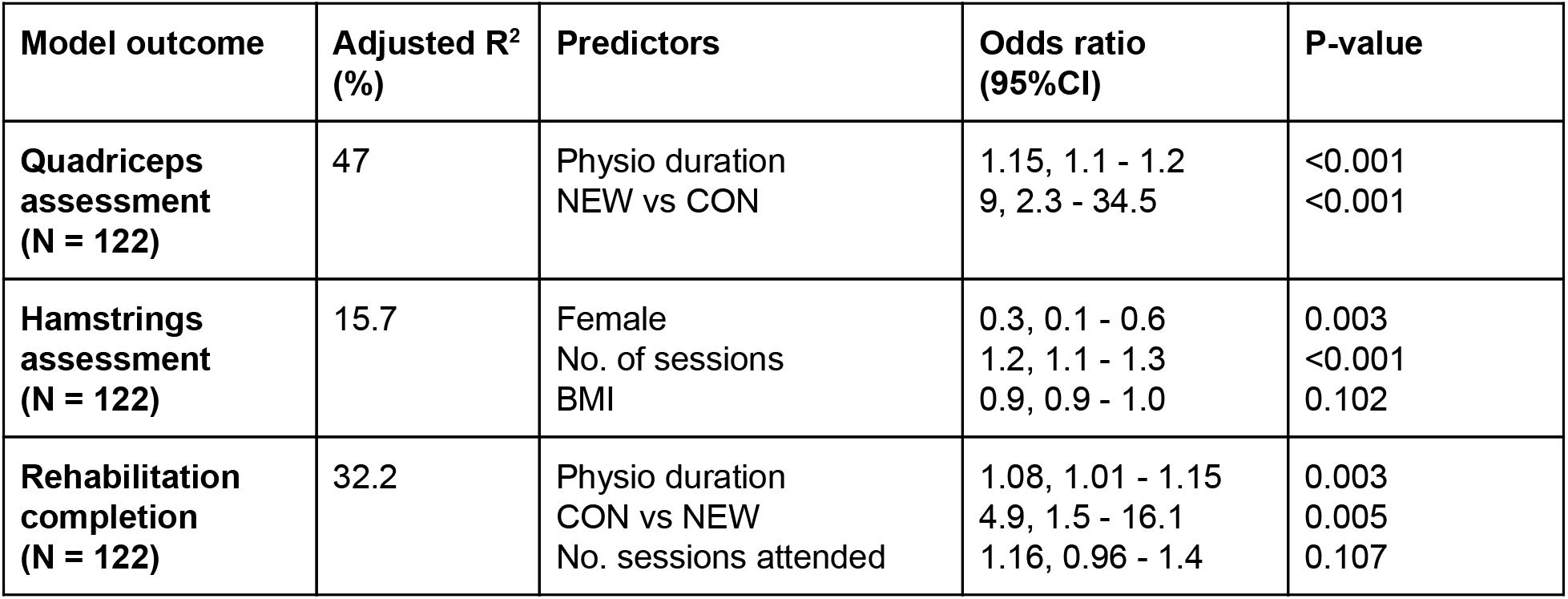
Summary of logistic regression results for assessment and patient outcomes, with adjusted odds ratio and 95% confidence intervals.

| **Model outcome** | **Adjusted R<sup>2</sup> (%)** | **Predictors** | **Odds ratio (95%CI)** | **P-value** |
| --- | --- | --- | --- | --- |
| **Quadriceps assessment (N = 122)** | 47 | Physio duration<br>NEW vs CON | 1.15, 1.1 - 1.2<br>9, 2.3 - 34.5 | <0.001<br><0.001 |
| **Hamstrings assessment (N = 122)** | 15.7 | Female<br>No. of sessions<br>BMI | 0.3, 0.1 - 0.6<br>1.2, 1.1 - 1.3<br>0.9, 0.9 - 1.0 | 0.003<br><0.001<br>0.102 |
| **Rehabilitation completion (N = 122)** | 32.2 | Physio duration<br>CON vs NEW<br>No. sessions attended | 1.08, 1.01 - 1.15<br>4.9, 1.5 - 16.1<br>1.16, 0.96 - 1.4 | 0.003<br>0.005<br>0.107 |

## Discussion

Our results suggest that a phase-based progressive rehabilitation program leads to increased utilisation of some evidence-based outcome measures by treating physiotherapists. A phase-based model also appears to influence certain service metrics, resulting in a longer duration of physiotherapy rehabilitation and a higher number of attended physiotherapy sessions. It does not appear to increase rehabilitation completion rates, or rates of return to sport and activity. Following the introduction of a phase-based model of care, the incidence of physiotherapists using objective outcome measures significantly increased for strength assessment of quadriceps and kinetic chain strength (including calf, glute or trunk) and single leg squat performance. This is a positive finding suggestive that the NEW model of care increased compliance with evidence-based guidelines. The importance of monitoring and increasing lower limb muscle strength, particularly quadriceps, in rehabilitation post ACLR is well accepted within current literature ^1,12,14,21^.

There is currently no ideal duration or dose of physiotherapy identified in the literature ^9,13,16,21^. In this study, ‘completion’ of physiotherapy indicated that the treating physiotherapist discharged the patient from the service. This would generally indicate that the patient is meeting certain physical outcomes (such as range of motion, strength and functional measurements) and has been given return to activity or sport advice. In comparison, being discharged due to failing to attend could occur at any time point post-surgery, and therefore at varied stages of rehabilitation. This study showed that attending physiotherapy for a longer duration was significantly associated with higher rehabilitation ‘completion’ rates. Interestingly, patients in the CONTROL group were more likely to complete rehabilitation compared to the NEW group. A potential explanation may be that prior to the new model of care there was less guidance around discharge criteria, relying on the individual clinicians’ judgement and experience to make that decision. This may have led to premature discharge and return to activity. In comparison, a phase-based model where clinicians are utilising objective outcomes to guide decision making may result in lower rates of discharge by the physiotherapist due to patients not achieving recommended criteria.

Follow-up phone calls investigated reasons why patients may fail to attend appointments, and subsequently be discharged from the service. Two of the most common reasons identified were *‘Thought they were discharged’* and ‘*happy with knee*’. Both these reasons indicate a potential disconnect between patient and physiotherapist expectations. In some cases, physiotherapists may be striving to achieve rehabilitation goals based on existing literature in sporting populations; however, not all public system ACLR patients may share these goals. The public hospital ACLR cohort includes a number of patients with poor pre-injury strength and conditioning, with little or no desire to return to sport. This introduces difficulty in establishing clear functional goals, and patients can experience low motivation to participate in rehabilitation. This may contribute to them ceasing rehabilitation sooner, and therefore not ‘completing’ physiotherapy. Patients participating in lower level, social sport can also demonstrate poor movement quality, a potential contributing factor to them sustaining their initial injury. This presents challenges when physiotherapists work towards achievement of movement-quality based rehabilitation criteria recommended in return to sport literature. Many of the public setting patient cohort will never need to achieve these criteria to return to their desired level of function; however, not achieving these criteria may elicit concern for treating physiotherapists.

Regardless of the model of care, poor attendance rates, low rehabilitation completion rates and reduced functional outcomes were observed. This highlights the importance of both patient selection for surgery, and the need for standardised education from all clinicians (surgeons and physiotherapists) regarding the intense rehabilitation requirements post ACLR. Heightening patients’ expectations may contribute to increased motivation and improved attendance ^10^. Given the literature reports satisfactory function can be achieved with conservative management ^10,11^, it is important this option is explored in those patients who may not have a desire to return to sport. The low rehabilitation completion rates seen in both groups may also be indicative of a lack of patient engagement in ACLR rehabilitation programs. Exploring patient factors that contribute to rehabilitation adherence and engagement would empower public services to develop potential strategies to address this issue.

Although this study has provided new information regarding the implications of new models of care in an ACL rehabilitation setting, its limitations should also be recognised. The retrospective, observational design precludes the determination of causal relationships between our findings and the model of care change. Additionally, some participants engaged in rehabilitation across both models due to the use of a specific cut-off date in group allocation. It should also be recognised that data collection was not blinded, and the researcher was involved in the service development and data collection. Data were captured from clinical notes that were not specifically documented for research purposes, and documentation was occasionally unclear and required interpretation, contributing to potential misclassification bias. When collecting data pertaining to physiotherapist use of outcome measures, if a physiotherapist had assessed that outcome measure at least once in their rehabilitation it was considered compliant, potentially leading to an over-representation of outcome measure use. A low response rate to the phone call follow-ups also made it difficult to extrapolate statistical relationships between the model of care change and return to sport or activity rates. Lastly, this study collected data on physiotherapy utilisation of outcome measures, however did not report on specific clinical patient outcomes. A prospective study investigating whether rehabilitation attendance and duration in a public hospital setting impacts the achievement of certain evidence-based outcomes (e.g. muscle strength, hop tests) would help inform ALCR rehabilitation models of care.

## Conclusion

This study determined that in patients electing to undergo supervised rehabilitation in a physiotherapy department in a public hospital, a new model of care incorporating a phase-based rehabilitation program achieved the following; an increase in physiotherapist utilisation of evidence-based outcome measures, namely, quadriceps strength, kinetic chain muscle strength, and single leg squat performance; an increased total duration of rehabilitation and an increased total number of physiotherapy sessions attended. Despite this, rehabilitation completion rates decreased with the new model of care. Further work is required to understand the relationships between rehabilitation models of care and key patient metrics, particularly functional outcome.

## Data Availability

Blinded data will be available on reasonable request.

## Acknowledgements

The authors wish to acknowledge the contribution of Loren Barton, for her assistance with chart review and the introduction of the new model of care into the department.

